# Temporal analysis of the clinical evolution of confirmed cases of COVID-19 in the state of Mato Grosso do Sul - Brazil

**DOI:** 10.1101/2020.09.21.20198812

**Authors:** Carolina Mariano Pompeo, Marcos Antonio Ferreira, Andréia Insabralde de Queiroz Cardoso, Luciana Scarlazzari Costa, Mercy da Costa Souza, Felipe Machado Mota, Maria Lúcia Ivo

## Abstract

The objective was to analyze the evolution of confirmed cases of COVID-19 in the first four months of the pandemic in Mato Grosso do Sul, a state in the Center-West region of Brazil, as well as the factors related to the prevalence of deaths. This was an observational study with a cross-sectional and time series design based on data from the information system of the State Department of Health of Mato Grosso do Sul, Brazil. The microdata from the epidemiological bulletin is open and in the public domain; consultation was carried out from March to July 2020. The incidences were stratified per 100,000 inhabitants. The cross-section study was conducted to describe COVID-19 cases, and the trend analysis was performed using polynomial regression models for time series, with R-Studio software and a significance level of 5%. There was a predominance of women among the cases, and of men in terms of deaths. The presence of comorbidities was statistically related to mortality, particularly lung disease and diabetes, and the mean age of the deaths was 67.7 years. Even though the macro-region of the state capital, Campo Grande, had a higher number of cases, the most fatalities were in the macro-region of Corumbá. The trend curve demonstrated discreet growth in the incidence of cases between epidemiological weeks 11 and 19, with a significant increase in week 20 throughout the state. The trend for COVID-19 in the state of Mato Grosso do Sul was upward and regular, but there was an important and alarming exponential increase. The health authorities should adopt the necessary measures to enforce health precautions and encourage social distancing of the population so that health services will be able to care for those afflicted by the disease, especially older people, those with comorbidities, and vulnerable sectors of the population.

## Introduction

The respiratory disease known as *Coronavirus Disease* 2019 (COVID-19) is caused by the virus *Severe Acute Respiratory Syndrome, Coronavirus 2* (SARS-CoV-2)^1^. CoVs are from the order *Nidovirales*, family *Coronaviridae* and subfamily *Coronavirinae*. They are single-stranded RNA virus groups that can cause respiratory, liver, gastrointestinal and neurological diseases in animal species and humans^2^.

Historically, in 2003, the first major epidemic of the 21^st^ century arose in China, caused by the virus *Severe Acute Respiratory Syndrome, Coronavirus* (SARS-CoV), subtypes of the virus *Influenza A*, which have also sparked epidemics around the world. In 2014, the variation H5N1 was responsible for an epidemic in Asian countries and, in 2009, the first pandemic of the 21^st^ century occurred, caused by another variation of the virus, H1N1. In 2012, the *Middle East Respiratory Syndrome Coronavirus* (MERS-CoV) gave rise to an epidemic that started in Saudi Arabia and spread to another 27 countries^2-4^.

COVID-19 was first detected at the end of December 2019, in the city of Wuhan in China, and was declared an international public health emergency on January 30, 2020. This marked the third highly pathogenic introduction of the coronavirus in humans in the 21^st^ century^1^.

In Mato Grosso do Sul, a state located in the Center-West region of Brazil, the first cases of COVID-19 were identified in the beginning of March 2020, and the first death occurred on the 31^st^ of that same month. The first two confirmations were people who had had contact with positive cases originating from the states of São Paulo and Rio de Janeiro. The first death in the state was a 64-year-old woman infected by the coronavirus after having had contact with family members coming from Belgium^5,6^. On April 13, the first case of community transmission in the state was confirmed, and the number of cases has substantially increased since then^7^.

Various measures were recommended by the World Health Organization for countries around the world to reduce increased dissemination of the COVID-19 virus^8^. The transmission of respiratory pathogens, such as the coronavirus, depends on their being transported by secretions transmitted between people in the form of aerosols, droplets, or secretions, or by direct contact with mucosa^1,9^.

The mortality rate for COVID-19 has still not been clearly determined, since it is rising in most countries. However, it has already been proven that people with comorbidities, such as cardiovascular diseases, diabetes, cancers and immunosuppression, in addition to older people, are at greater risk. The contagion rate seems to vary among countries, which may be directly related to local health responses for social isolation and care of the affected population^10^.

The rapid dynamics of the COVID-19 pandemic are a challenge and source of concern for healthcare systems in different locations, especially emerging countries such as Brazil, because fragile social and economic contexts may lead to underreporting and make adequate management of the pandemic more difficult, which increases uncertainties as to the dynamics of the disease^11^.

Despite the implementation of measures and strategies to reduce the transmission of COVID-19, the number of cases is expected to rise in all the Brazilian states. There is a concern about the availability of intensive care unit beds and ventilator support for hospitalized patients, in addition to specific diagnostic tests in sufficient amounts to monitor the development of the disease^12^.

Due to the epidemiological importance of COVID-19 for Brazil and the world, it is necessary to ascertain the real situation of the pandemic in the Brazilian states in order to provide support for the creation of guidelines and the decisions that need to be made. Therefore, this study sought to analyze the temporal trend of confirmed cases of COVID-19 in the first four months of the pandemic in Mato Grosso do Sul, a state in the Center-West region of Brazil, and in its macro-regions, as well as the factors related to the prevalence of deaths.

## Materials and Methods

This was an observational, time series study of confirmed cases of COVID-19 in Mato Grosso do Sul, Brazil, in the first four months of the pandemic – a period that spans the epidemiological weeks (EW) 11 to 28. The data was from an open database of the State Department of Health of Mato Grosso, available in microdata from the COVID-19 epidemiological bulletin. The population for calculating the incidence was obtained through data from the Brazilian Institute of Geography and Statistics (IBGE) from 2019. Both databases are in the public domain and do not require previous consideration or authorization by a research ethics committee. The data was collected between March and July 2020 and was tabulated on spreadsheets using Microsoft Excel 2010^®^.

In the statistical analysis, a cross-sectional study was first conducted with data from COVID-19 patients from the state of Mato Grosso do Sul that sought to describe the COVID-19 cases in the state according to situation/status of the disease and characteristics of the population.

A regression model was estimated to verify factors related to death/non-death, which in this case was considered as a dependent variable. The situation of the patients originally coded in the database as death, non-death, recovered or in home treatment were treated in a grouped manner for analysis purposes. Therefore two groups were created – death and non-death – with the latter including those who recovered and those in home treatment. The independent variables were sociodemographics, comorbidities, and signs and symptoms of the disease. A total of 555 patients was excluded from the sample, whose information was ignored, and 37 patients who were hospitalized.

The data was initially presented through descriptive statistics, absolute values, percentage, and mean, minimum and maximum values, according to the type of variable. Association tests were then performed using the chi-square or Fisher’s exact test and the Student’s t-test for comparison of means. Following this, the risk measurement and crude prevalence ratio were estimated, through univariate Poisson modeling, to assess the magnitude of the risk of death based on the characteristics of the study.

Last, the Poisson multiple regression model to verify the factors related to the prevalence of death among patients with COVID-19 was adjusted. In this model, the possible confounding variables were determined and possible interactions were tested (respiratory difficulties, pulmonary problems, sex and race).

Apart from the regression analysis, a trend analysis was performed in relation to EW 11 to 28 using polynomial regression models for time series. The dependent variable refers to the COVID-19 incidence coefficients and the independent variable refers to the epidemiological weeks. Temporal graphs were built for the coefficients, for the state and macro-regions, to verify the possible format of the trend curve to be studied. Next, polynomial regression models were estimated; first, the linear regression model was tested (*Y* = *β*_0_ + *β*_1_*X*) and then the more complex models, such as those of the 2^nd^ order (*Y* = *β*_0_ + *β*_1_ *X* + *β*_1_ *X*^2^), 3rd order (*Y* = *β*_0_ + *β*_1_*X* + *β*_1_*X*^2^ + *β*_1_*X*^3^) and exponential.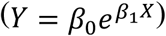.

The time variable (EW) was centralized to the mean point of the series to avoid auto-correlations. The models were chosen according to their statistical significance, residual analysis, and coefficient of determination (R^2^).

R-Studio statistical software with a significance level of 5% was used for all the analyses.

## Results

From the first confirmed case of COVID-19 in EW 11 until the end of EW 28, a total of 14,092 cases was diagnosed, with an incidence of 507.09 per 100,000 inhabitants. The macro-region of Campo Grande, the most populous in the state, which is home to the state capital, had the highest number of confirmations of the disease (48%). However, the highest incidence was in the macro-region of Dourados with 688.24 cases per 100,000 inhabitants. The second highest incidence occurred in Corumbá (488.96), followed by Campo Grande (445.45) and then Três Lagoas (309.21). Among those diagnosed, approximately 93% were asymptomatic patients.

There was a higher number of infections among young adults in the age group of 30 to 39 years old (27.5%), and the highest number of deaths occurred among older people (above 60 years old) in more than 70% of the case. In all the age groups, most patients recovered from the disease. Of the total number of deaths, more than 85% of the cases had at least one associated comorbidity, and almost half of the cases (48%) had two or more. The distribution of the cases according to health situation and age is presented in Figure 1.

**Figure 1:**
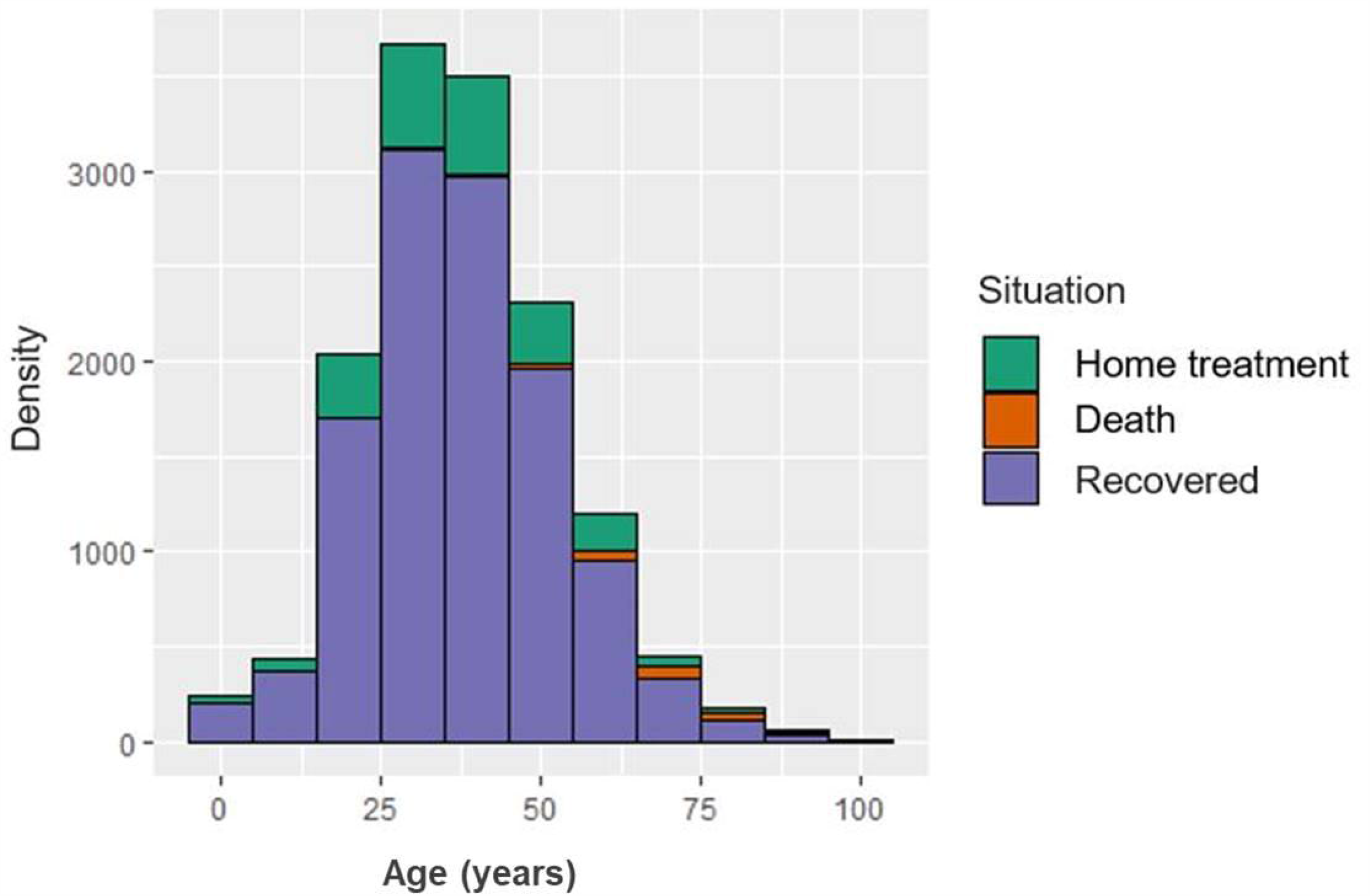
Distribution by age and situation of patients with COVID-19 in Mato Grosso do Sul, Brazil, 2020.

Among the state’s macro-regions, the highest fatality rate was 2.58 in Corumbá, followed by 2.17 in Três Lagoas and 1.66 in Dourados; the lowest fatality rate was in the macro-region of Campo Grande.

The variables corresponding to age, ethnic group, sex, macro-region, race, fever, sore throat, respiratory difficulties, number of symptoms, type of test for detecting COVID-19, pulmonary problems, diabetes, number of comorbidities, and occupation (whether a health professional or not) had a statistical relationship with the patients’ situation. The groups of patients that died, and those that did not, were heterogeneous in relation to these characteristics. The breakdown of the data is presented in Table 1.

**Table 1.** Characteristics of the patients with confirmed cases of infection by COVID-19 in Mato Grosso do Sul, SES/MS^1^. Brazil, 2020 (n=14,092).

| Characteristics | Total<br>(n=14,092) | Death<br>(n=221) | Non-death<br>(n=13,871) | p-value |
| --- | --- | --- | --- | --- |
| **Mean age (min-max)** | 38.4 (0.0 - 102.0) | 67.7 (25.0 - 100.0) | 37.9 (0.0 - 102.0) | **< 0.001<sup>2</sup>** |
| **Age groups, n (%)** |  |  |  | **< 0.001** |
| 0 – 9 years old | 404 (2.9) | 0 (0.0) | 404 (2.9) |  |
| 10 – 19 years old | 716 (5.1) | 0 (0.0) | 716 (5.2) |  |
| 20 – 29 years old | 2,975 (21.1) | 4 (1.8) | 2,971 (21.4) |  |
| 30 – 39 years old | 3,877 (27.5) | 9 (4.1) | 3,868 (27.9) |  |
| 40 – 49 years old | 2,981 (21.1) | 12 (5.4) | 2,969 (21.4) |  |
| 50 – 59 years old | 1,879 (13.3) | 35 (15.8) | 1,844 (13.3) |  |
| 60 years old or more | 1,260 (8.9) | 161 (72.9) | 1,099 (7.9) |  |
| **Sex, n (%)** |  |  |  | **0.009** |
| Male | 6,719 (47.7) | 125 (56.6) | 6,594 (47.5) |  |
| Female | 7,373 (52.3) | 96 (43.4) | 7,277 (52.5) |  |
| **Macro-region, n (%)** |  |  |  | **0.022** |
| Campo Grande | 6,771 (48.0) | 89 (40.3) | 6,682 (48.2) |  |
| Corumbá | 659 (4.7) | 17 (7.7) | 642 (4.6) |  |
| Dourados | 5,785 (41.1) | 96 (43.4) | 5,689 (41.0) |  |
| Três Lagoas | 877 (6.2) | 19 (8.6) | 858 (6.2) |  |
| **Race/color** |  |  |  | **0.019** |
| Yellow | 953 (7.2) | 7 (3.6) | 946 (7.3) |  |
| White | 7,204 (54.5) | 94 (47.7) | 7,110 (54.6) |  |
| Indigenous | 278 (2.1) | 4 (2.0) | 274 (2.1) |  |
| Brown | 4,302 (32.5) | 84 (42.6) | 4,218 (32.4) |  |
| Black | 487 (3.7) | 8 (4.1) | 479 (3.6) |  |
| **Signs/Symptoms, n (%)** |  |  |  |  |
| Fever |  |  |  |  |
| Yes | 781 (78.5) | 138 (66.7) | 643 (81.6) | **<0.001** |
| No | 214 (21.5) | 69 (33.3) | 145 (18.4) |  |
| Cough |  |  |  |  |
| Yes | 849 (84.1) | 168 (80.8) | 681 (85.1) | 0.165 |
| No | 160 (15.8) | 40 (19.2) | 120 (14.9) |  |
| Sore throat |  |  |  |  |
| Yes | 347 (36.7) | 54 (28.0) | 293 (38.9) | **0.006** |
| No | 599 (63.3) | 139 (72.0) | 460 (61.1) |  |
| Difficulty breathing |  |  |  |  |
| Yes | 663 (67.5) | 157 (77.0) | 506 (65.0) | **0.001** |
| No | 319 (32.5) | 47 (23.0) | 272 (35.0) |  |
| Number of symptoms |  |  |  |  |
| None | 13,078 (92.8) | 10 (4.5) | 13,068 (94.2) | **<0.001** |
| One or more | 1,014 (7.2) | 211 (95.5) | 803 (5.8) |  |
| **Type of test** |  |  |  |  |
| RT-PCR | 8,087 (57.6) | 199 (90.0) | 7,888 (57.1) | **<0.001** |
| Serology | 135 (0.0) | 0 (0.0) | 135 (1.0) |  |
| Quick test | 5,806 (41.4) | 22 (10.0) | 5,784 (41.9) |  |
| **Comorbidities, n (%)** |  |  |  |  |
| Pulmonary problem |  |  |  |  |
| Yes | 679 (71.1) | 153 (77.3) | 526 (69.5) | **0.040** |
| No | 276 (28.9) | 45 (22.7) | 231 (30.5) |  |
| Kidney problem |  |  |  |  |
| Yes | 6 (1.8) | 3 (2.9) | 3 (1.3) | 0.376 |
| No | 330 (98.2) | 100 (97.1) | 230 (98.7) |  |
| Weak immunity |  |  |  |  |
| Yes | 2 (0.0) | 0 (0.0) | 2 (0.9) | ~1.000 |
| No | 333 (99.4) | 103 (100.0) | 230 (99.1) |  |
| Diabetes |  |  |  |  |
| Yes | 257 (58.7) | 90 (66.7) | 167 (55.1) | **0.030** |
| No | 181 (41.3) | 45 (33.3) | 136 (44.9) |  |
| Immunosuppression |  |  |  |  |
| Yes | 23 (6.6) | 8 (7.5) | 15 (6.2) | 0.841 |
| No | 325 (93.4) | 99 (92.5) | 226 (93.8) |  |
| Heart disease |  |  |  |  |
| Yes | 236 (54.0) | 82 (61.2) | 154 (50.8) | 0.060 |
| No | 201 (46.0) | 52 (38.8) | 149 (49.2) |  |
| High-risk pregnancy |  |  |  |  |
| Yes | 6 (1.7) | 2 (2.7) | 4 (1.4) | 0.607 |
| No | 355 (98.3) | 72 (97.3) | 283 (98.6) |  |
| Number of comorbidities |  |  |  |  |
| None | 13,290 (94.3) | 32 (14.5) | 13,258 (95.6) | **<0.001** |
| One | 494 (3.5) | 83 (37.5) | 411 (2.9) |  |
| Two or more | 308 (2.2) | 106 (48.0) | 202 (1.5) |  |
| **Health professional, n (%)** |  |  |  |  |
| Yes | 1,835 (13.0) | 7 (3.2) | 1,828 (13.2) | **< 0.001** |
| No | 12,257 (86.9) | 214 (86.8) | 12,043 (86.8) |  |
| **Security team member,** |  |  |  |  |
| **n (%)** |  |  |  |  |
| Yes | 198 (1.6) | 0 (0.0) | 198 (1.6) | ~1.000 |
| No | 12,472 (98.4) | 1 (0.0) | 12,471 (98.4) |  |

<sup>1</sup> SES/MS: State Department of Health of Mato Grosso do Sul
<sup>2</sup> Mean comparison test (Student's test)

The characteristics that had statistically significant associations with the patient’s situation in the multiple context were age, classified into distribution quartiles, location of the patient (macro-region), and number of comorbidities, controlled by sex and race/color. The interactions between respiratory difficulties and pulmonary problems and between sex and race/color were tested, but did not reach statistical significance.

In relation to the factors associated with prevalence of death by COVID-19, the prevalence was 13.34 times higher in the age group of 60 years old or more, compared to the age group of 0 to 29 years (baseline), regardless of location and number of comorbidities. Individuals with two or more associated comorbidities had 62.37 times more risk of dying, compared to individuals without comorbidities, regardless of location and age group (Table 2).

**Table 2.** Factors associated with prevalence of death by COVID-19 in patients with confirmed cases in Mato Grosso do Sul, SES/MS^1^. Brazil, 2020 (n=14.092).

| Characteristics | $\beta_{\text{crude}}$ (CI 95%) | $PR_{\text{crude}}$ (CI 95%) | $PR^2_{\text{adj}}$ (CI 95%) |
| --- | --- | --- | --- |
| **Age, quartiles (years old)** |  |  |  |
| 00 – 29 |  | Baseline |  |
| 30 – 49 | 1.14 (0.18 - 2.37) | 3.13 (1.19 - 10.74) |  |
| 50 - 59 | 2.94 (2.03 - 4.16) | 19.07 (7.62 - 63.82) | 5.24 (2.02 - 17.89) |
| 60 or more | 4.87 (4.01 - 6.05) | 130.82 (55.39 - 425.05) | 13.34 (5.36 - 33.66) |
| **Macro-region** |  |  |  |
| Campo Grande |  | Baseline |  |
| Corumbá | 0.67 (0.12 - 1.16) | 1.96 (1.12 - 3.21) | 2.46 (1.39 - 4.08) |
| Dourados | 0.23 (-0.05 - 0.52) | 1.26 (0.94 - 1.68) | 1.41 (1.03 - 1.93) |
| Três Lagoas | 0.49 (-0.02 - 0.97) | 1.65 (0.97 - 2.64) | 1.81 (1.03 - 3.02) |
| **Number of comorbidities** |  |  |  |
| None |  | Baseline |  |
| One | 4.24 (3.85 - 4.67) | 69.78 (46.91 - 106.38) | 42.73 (26.43 - 71.63) |
| Two or more | 4.96 (4.58 - 5.37) | 142.93 (97.48 - 215.54) | 62.37 (38.52 - 105.06) |

<sup>1</sup> SES/MS: State Department of Health of Mato Grosso do Sul
<sup>2</sup> Prevalence Ratio obtained through the Poisson regression model, adjusted by sex and race

Figure 2 presents the growth curves of the COVID-19 incidence coefficients in Mato Grosso do Sul and the state’s macro-regions, in the period encompassing epidemiological weeks 11 to 18. Growth was similar among the regions until week 20, after which time the curves were different between the regions, particularly the Dourados region, which had higher incidences from that time on and then dropped off at week 26.

**Figure 2:**
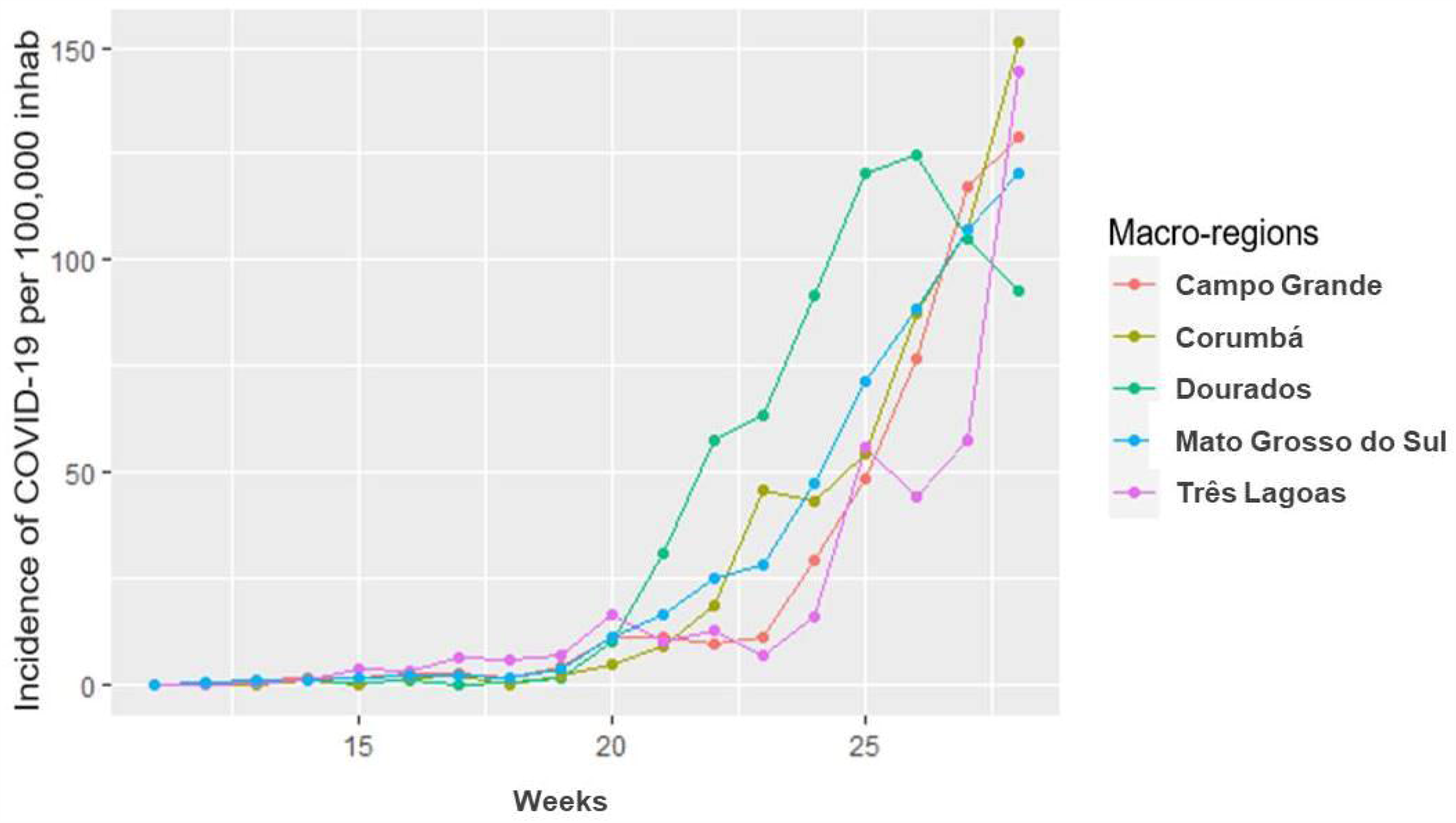
Evolution of cases of COVID-19, by macro-region and epidemiological week.

The overall coefficient for incidence and death remained stable and had similar curves until week 19. Starting at week 20, a distance opened up between them, with an increase in the overall incidence coefficient (Figure 3).

**Figure 3:**
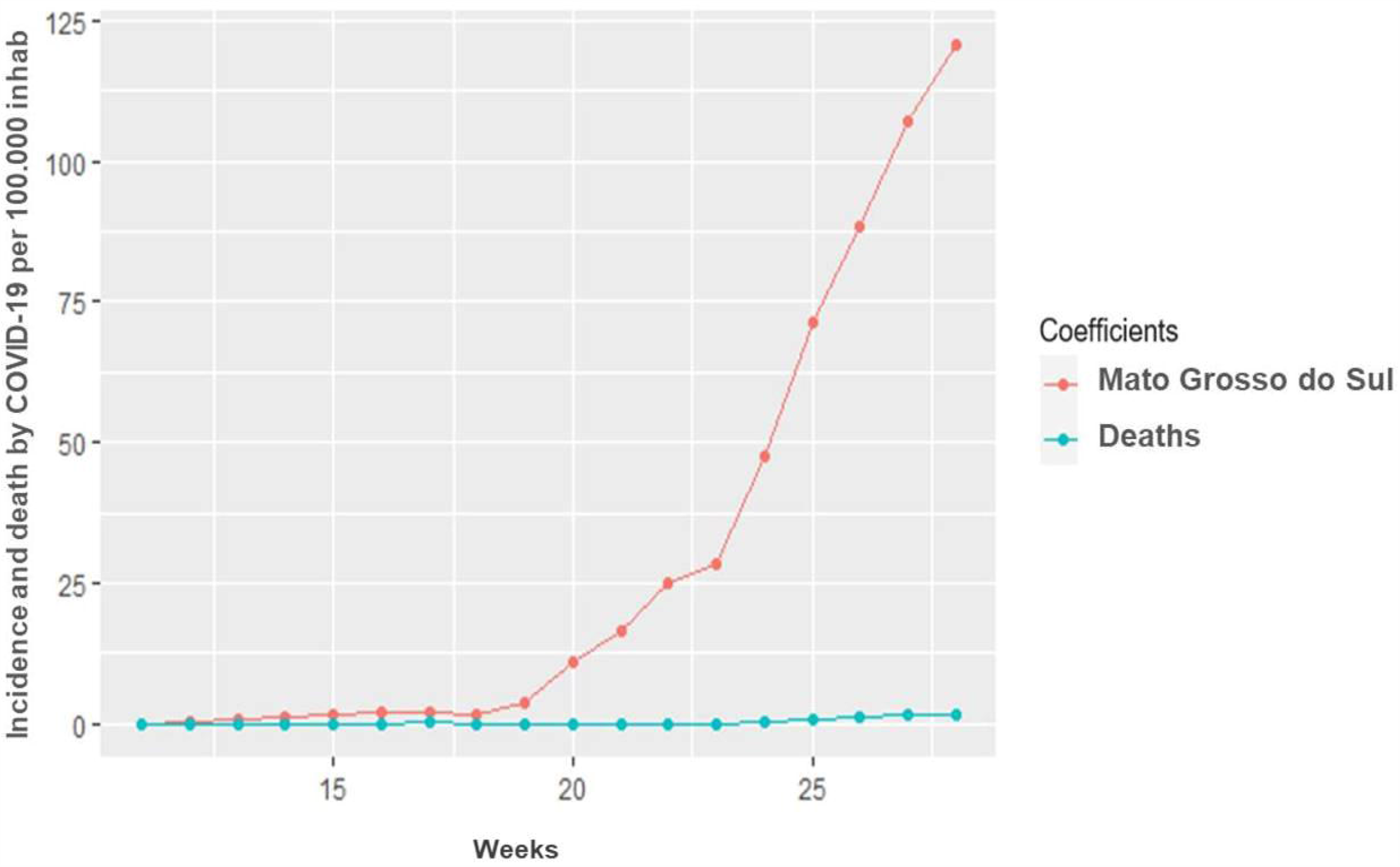
Evolution of cases and deaths by COVID-19, in Mato Grosso do Sul, Brazil.

Table 3 and Figure 4 present the results of trend analysis for COVID-19 incidence coefficients in the state of Mato Grosso do Sul and its macro-regions for EW 11 to 28. All the models were statistically significant with coefficients of determinants over 0.85.

**Table 3.**
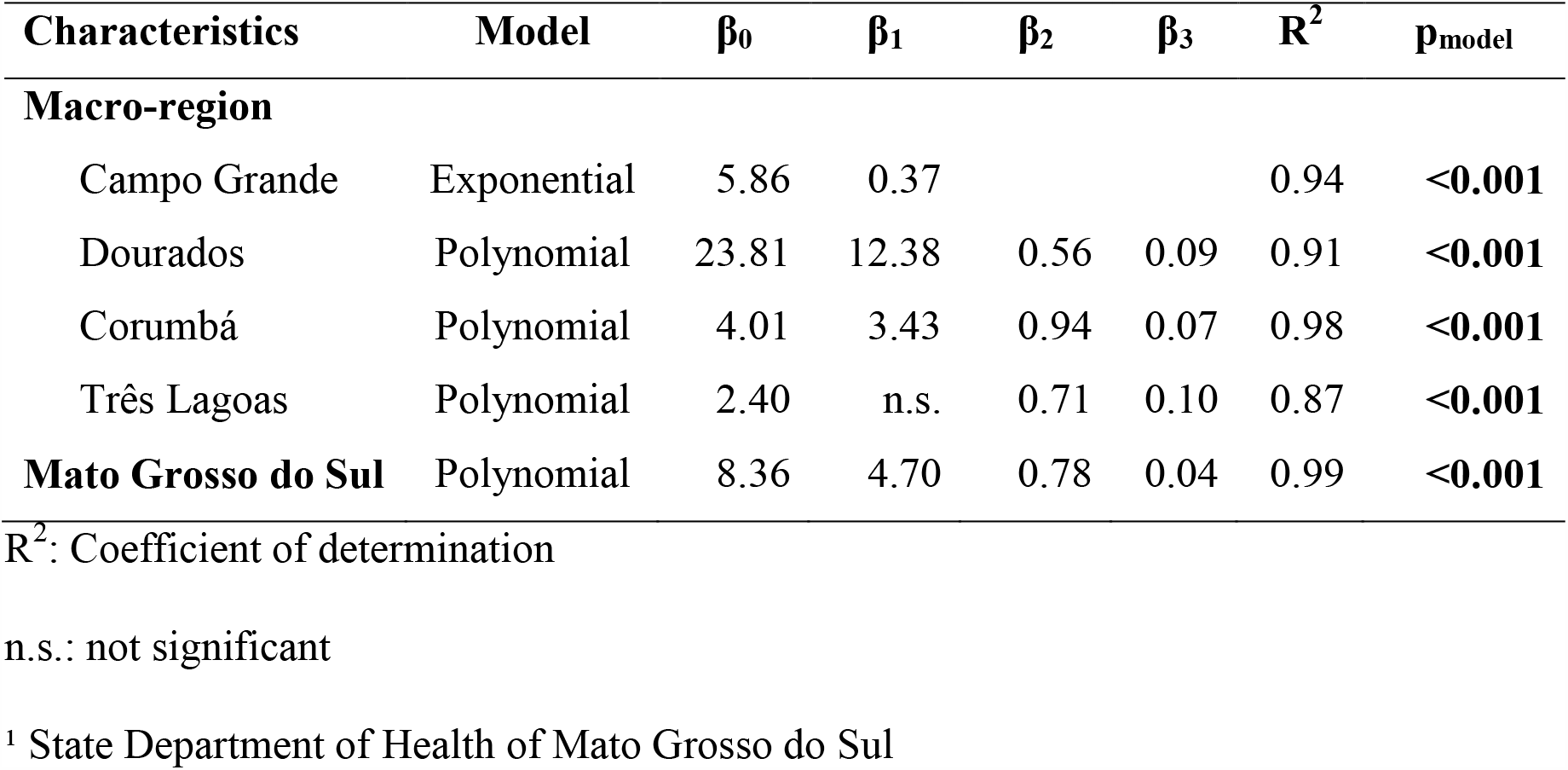
Analysis of the trend in the number of cases of COVID-19 according to macro-region, micro-region, sex, age group and comorbidities, SES/MS^1^. Brazil, 2020 (n=14.092).

**Figure 4:**
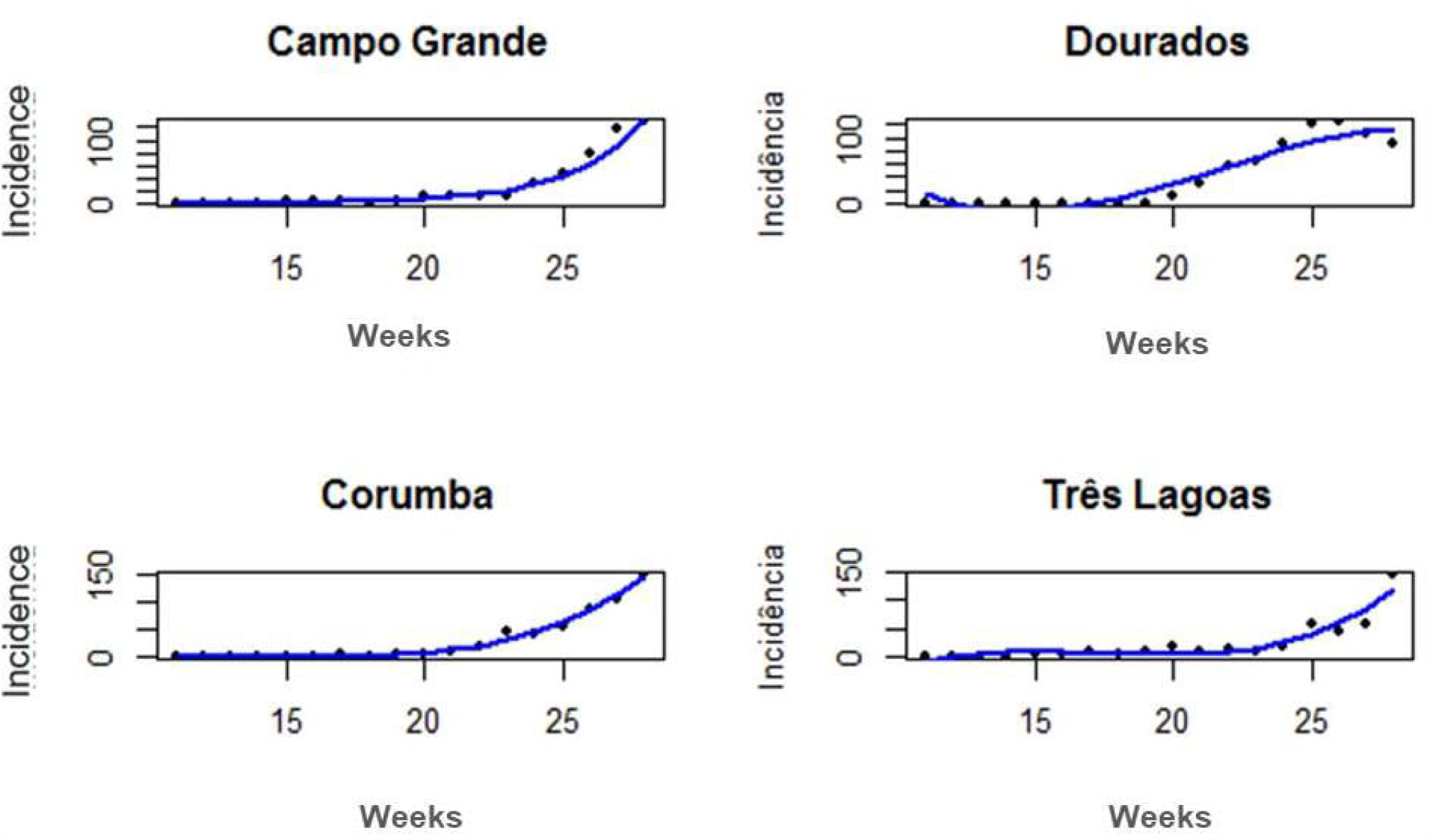
Values observed and estimated by the trend models for COVID-19 incidence coefficients in the macro-regions of Mato Grosso do Sul, Brazil.

## Discussion

This study examined the incidence, characteristics and temporal evolution of COVID-19 cases in Mato Grosso do Sul, Brazil, in epidemiological weeks 11 to 28. Even though the data indicated a substantial number of recovered cases, among the more serious cases of the disease, 222 (1.57%) ended in death. Of these, the majority had at least one associated comorbidity or involved people over 60 years old, with or without associated chronic diseases^13,14^.

In addition, the presence of one and, mainly, two or more comorbidities were independent risk factors for death, similar to that observed in the international context of the pandemic^15-17^

Among the comorbidities found, pulmonary disease and diabetes had an association with the occurrence of deaths, which is consistent with increased risk of mortality in patients with these comorbidities observed in other studies^17,18^.

There was a higher number of deaths among men, even though the incidence of cases was higher among women. Studies conducted during this pandemic and in other SARS pandemics have also indicated more serious evolution of the disease among men and associations with more deaths among these cases^18,19.^ The higher fatality rate noted among the brown race was also confirmed in Brazil’s North region, where this rate was found to be higher among brown and black individuals, which may be related to differences in susceptibility to COVID-19 and access to adequate health care by this segment of the population^20,21^.

The signs and symptoms manifested by patients, such as fever and respiratory difficulties, were associated with higher mortality. The clinical manifestations of COVID-19 appear to vary from region to region^22^, although the most frequent ones in symptomatic patients have been found to be fever and fatigue, and the most common complications have been pneumonia and respiratory insufficiency^23,24^.

The higher distribution of the disease in the macro-region of Dourados may be due to population density, as well as the extensive territory of the region’s main city, which often results in the need to commute by public transportation, and the presence of work locations with large numbers of people on production lines, such as meatpacking plants - factors that favor dispersion and, consequently, viral contamination^25^. Of the most important cities in Mato Grosso do Sul, economically and population-wise, three stand out as at high risk for COVID-19 and account for approximately 44% of the state’s population: Campo Grande, Dourados and Três Lagoas^26^.

Although the incidence of cases was higher in the macro-region of Dourados, the largest proportion of deaths occurred in the region of Corumbá, which had a mortality rate of 2.58% and a statistically significant relationship among the regions. This index exceeded the calculated estimate for COVID-19 in China, where the mortality rate was 1.4%^27^, but was lower than the overall mortality rate of 6.52% in Mexico^28^. There are still many uncertainties and unknowns in relation to the viral mortality of COVID-19, since it is related to other factors^29^.

In terms of the evolution of cases, the data seems to suggest a growing and regular trend throughout the state, with a more significant increase after EW 20, the period that coincides with the start of winter in the Center-West region and, consequently, a period of higher transmission of respiratory diseases^30^.

There was also a statistically significant association in all the macro-regions of Mato Grosso do Sul, which enables the inference that the curve for COVID-19 is far from flattening, and may experience substantial and alarming growth. A mathematical model study conducted by researchers at the Federal University of Mato Grosso do Sul^31^ pointed out the exponential growth of the disease infection curve between the months of August and September, with probable collapse of the public health system if mitigation measures are not taken by managers^32^.

Understanding the dynamics of the disease and its ecological evidence and evolution worldwide are important factors for developing epidemiological perception and analysis of the impacts26,29. Therefore, this study sought to operate within this context, in order to contribute to a more complete picture of the epidemiological situation of COVID-19 in Brazil, and consequently in the world, in order to assist the scientific community and health managers with decision-making. One of the limitations of this study, similar to others with cross-sectional designs that use secondary data, is related to the time of the outcomes with regard to COVID-19 in the population, since there may be underestimation of real data due to underreporting and the presence of non-symptomatic carriers, in addition to the reality of the organization of local health services.

## Conclusion

The trend in relation to COVID-19 is growing and regular throughout the state of Mato Grosso do Sul, rising significantly after EW 20. This suggests that the state is far from flattening the curve, which has manifested exponential, substantial and alarming growth.

Health authorities must adopt appropriate measures to support health precautions and promote social distancing among the population in order to enable health services to care for those infected by the disease. Health actions need to especially target vulnerable groups and male patients due to their tendency toward higher mortality. Care should be reinforced among elderly people with comorbidities, as well as individuals in groups that are at risk of developing more serious forms of the disease.

## Data Availability

All data highlighted in the manuscript are available in an open database and contained in the text and tables

## Acknowledgment

The authors are grateful for the commitment and dedication of all professionals at the Maria Aparecida Pedrossian University Hospital of the Federal University of Mato Grosso do Sul in combating this serious crisis and to the employees of the State Department of Health of Mato Grosso do Sul - Brazil, for maintaining an open and comprehensive database for research.

